# Accuracy of a Second-Year Dermatology Resident’s Pre-Biopsy Diagnoses

**DOI:** 10.1101/2025.05.19.25327929

**Authors:** Emily M. Meara, Jessica K. Orofino, Vinod E. Nambudiri, Jeffrey S. Smith

## Abstract

Skin biopsies are heavily relied upon in dermatology to diagnose cutaneous malignancies. Mastering skin biopsies and recognizing skin cancer is a core component of dermatology residency training. However, limited data exists regarding evaluative standards for biopsy training and residents’ accuracy in diagnosing skin cancer. This study aimed to analyze pre- and post-biopsy diagnostic accuracy of a second-year resident in the Harvard Combined Dermatology Residency Program from July 2021 to June 2022. This resident recorded biopsies performed in specialty dermatology clinics across three large academic centers with attending oversight in addition to the hypothesized pre-biopsy diagnosis and the subsequent histopathologically confirmed diagnosis. Lesions were stratified into categories of squamous cell carcinoma (SCC), basal cell carcinoma (BCC), or benign lesions. Pre- and post-biopsy diagnoses were then compared. The resident correctly identified the diagnosis in 64% of cases and correctly classified all histologically confirmed skin cancers as SCC or BCC. The specificity for identifying an SCC or BCC was 56%. This study provides a preliminary foundational dataset for the development of evidence-based residency training guidelines to better prepare residents for independent clinical practice. Additionally, it serves as a model for providing trainees and programs with insight into variations in practice patterns.

## Introduction

Two of the most common reasons patients seek dermatologic care include skin examinations and evaluation of skin lesions. These visit types often rely on a dermatologist’s knowledge of cutaneous malignancies[1]. Accordingly, a core competency of dermatology residency training is the accurate diagnosis of skin cancers, including basal cell carcinoma (BCC) and squamous cell carcinoma (SCC). In the United States, approximately 3.6 million cases of BCCs and 1.8 million cases of SCCs are diagnosed annually, highlighting the importance of timely and accurate diagnosis[2].

Dermatology residents develop diagnostic proficiency through the integration of clinical history and visual inspection, including dermoscopy. While these techniques enhance diagnostic accuracy, they do not always result in complete concordance between pre-biopsy clinical impressions and post-biopsy histopathologic diagnoses. This discordance represents an opportunity to further investigate dermatology residency training, assessment, and optimization of biopsy utilization.

In the United States, several oversight organizations ensure that medical trainees receive adequate education in their chosen fields through established guidelines and training requirements. The American Board of Dermatology (ABD) is one such organization, outlining core competencies including the expectation that residents accurately identify concerning lesions requiring biopsy and perform a range of biopsy techniques[3]. This is further supported by the Accreditation Council for Graduate Medical Education (ACGME) guidelines, which emphasize resident responsibility and shared decision-making with patients regarding biopsy procedures.

As the field of dermatology evolves, guidelines are updated to reflect emerging needs and techniques, and ensuring that residents meet these evolving standards is a key responsibility of these oversight committees. Currently, the ACGME evaluates resident preparedness through competency logs, testing, and institution-level evaluation of milestones. However, there is no standardized approach to tracking the specific details of completed competencies, such as concordance between pre- and post-biopsy diagnoses. This study aims to help establish a foundation for evidence-based analysis of resident diagnostic accuracy over the course of training and to inform future approaches to residency evaluation.

One study tracking diagnostic concordance of dermatoscopic and dermatopathologic diagnoses of cutaneous malignancies from 2010 to 2019 found a significant increase in accuracy of SCC diagnosis from prior reports correlated with the use of a dermatoscope in evaluation[4]. This data was used to support the incorporation of dermoscopy into standard clinical practice. Such studies underscore the importance of systemic data collection in advancing evidence-based practice. This same study reported accuracy and specificity metrics for BCC and SCC diagnosis using dermoscopy, providing a benchmark for further research. Specificity for BCC was found to be 76.2% to 99%, with an accuracy of 98.5%, while SCC specificity was found to be 23.8% with an accuracy of 67.39%.

While dermatopathology remains the gold standard for diagnosing cutaneous malignancy, improved accuracy in resident’s initial assessment can facilitate more targeted utilization of biopsies, avoiding unnecessary risks while maximizing cancer detection. However, there remains a paucity of data regarding evaluative standards for biopsy training in residency and assessing resident accuracy in predicting and diagnosing cutaneous malignancies. With the shift towards competency-based curricula in dermatology training, there is a clear need for more data-driven guidelines. To address this gap, this pilot study analyzed a dataset of pre- and post-biopsy diagnoses collected across three large academic medical centers by one author (JSS) during his second year of dermatology residency training.

## Methods

This study is a retrospective review of 80 recorded skin biopsies performed by a second-year dermatology resident. These biopsies, including punch biopsies, shave biopsies, and excisions, were completed under attending supervision in specialty dermatology clinics from July 2021 to June 2022. The data was sourced from logs recorded by the resident. The logs recorded lesion location as well as whether the lesion had been identified by the patient or noted by the physician on physical exam. The pre-biopsy diagnosis was listed at the time of biopsy, and the post-biopsy diagnosis was listed following histopathologic confirmation by board-certified dermatopathologists. Data for each included biopsy contained a recorded pre-biopsy clinical impression with one primary diagnosis and one recorded post-biopsy final diagnosis. Data was excluded from analysis if pre-biopsy diagnosis was not limited to one diagnosis or was unclear, or if no singular post-biopsy diagnosis was listed. Concordance was characterized by identical pre- and post-biopsy designation into the stratified categories of BCC, SCC, or benign lesions. Concern for melanoma was limited and thus not included in this data. No IRB was required for this study, as no patient data was shared or recorded. Sample size limited data comparison between stratified groupings.

## Results

Of the 80 recorded biopsies where a reasonable concern for SCC or BCC was retained by the resident and/or attending, most biopsies were taken from the face (30%) and trunk (33%) (Figure 1A). Before biopsy, 54% of lesions were favored by the resident to be BCC, 18% as SCC, and 29% as benign lesions (Figure 1B). The post-biopsy histologic diagnosis breakdown identified 33% as BCC, 15% as SCC, and 53% as benign lesions (Figure 1C). Of all the biopsied lesions, the resident correctly identified the diagnosis in 64% of the cases and correctly diagnosed all histologically confirmed skin cancers as SCC or BCC, maintaining an overall specificity of 56% (Figure 2). The number needed to biopsy (NNB) was calculated as the total number of biopsies divided by the histologically diagnosed skin cancers, resulting in a value of 2.1.

**Figure 1:**
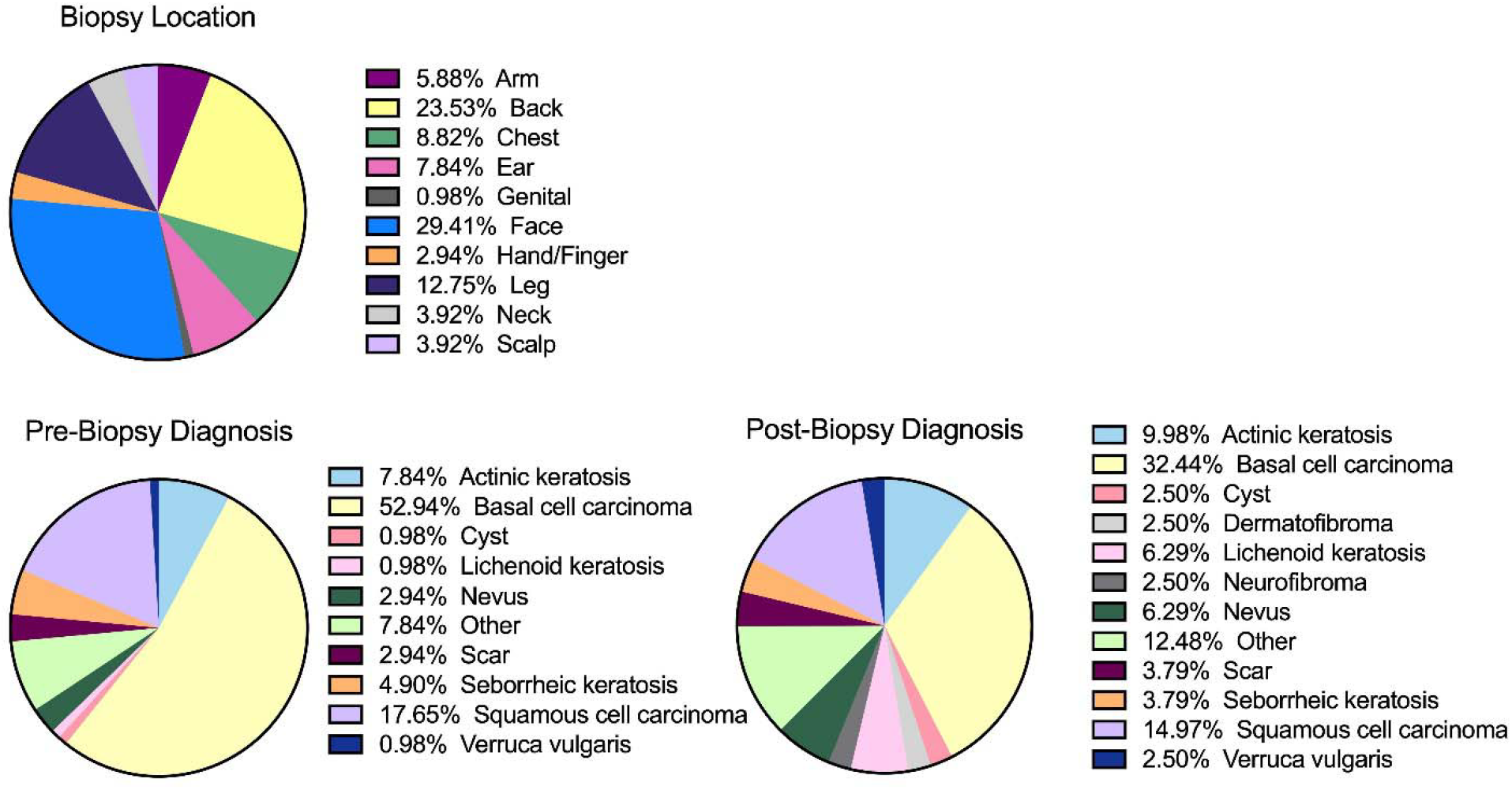
**A. Location of Biopsies.** Anatomical location of each recorded biopsy. **B. Pre-Biopsy Diagnosis**. Percentage of each diagnosis predicted by the resident prior to receiving the correlated histopathologic diagnosis. **C. Post-Biopsy Diagnosis**. Percentage of each diagnosis out of the total based on pathological determination from each biopsy sample.

**Figure 2:**
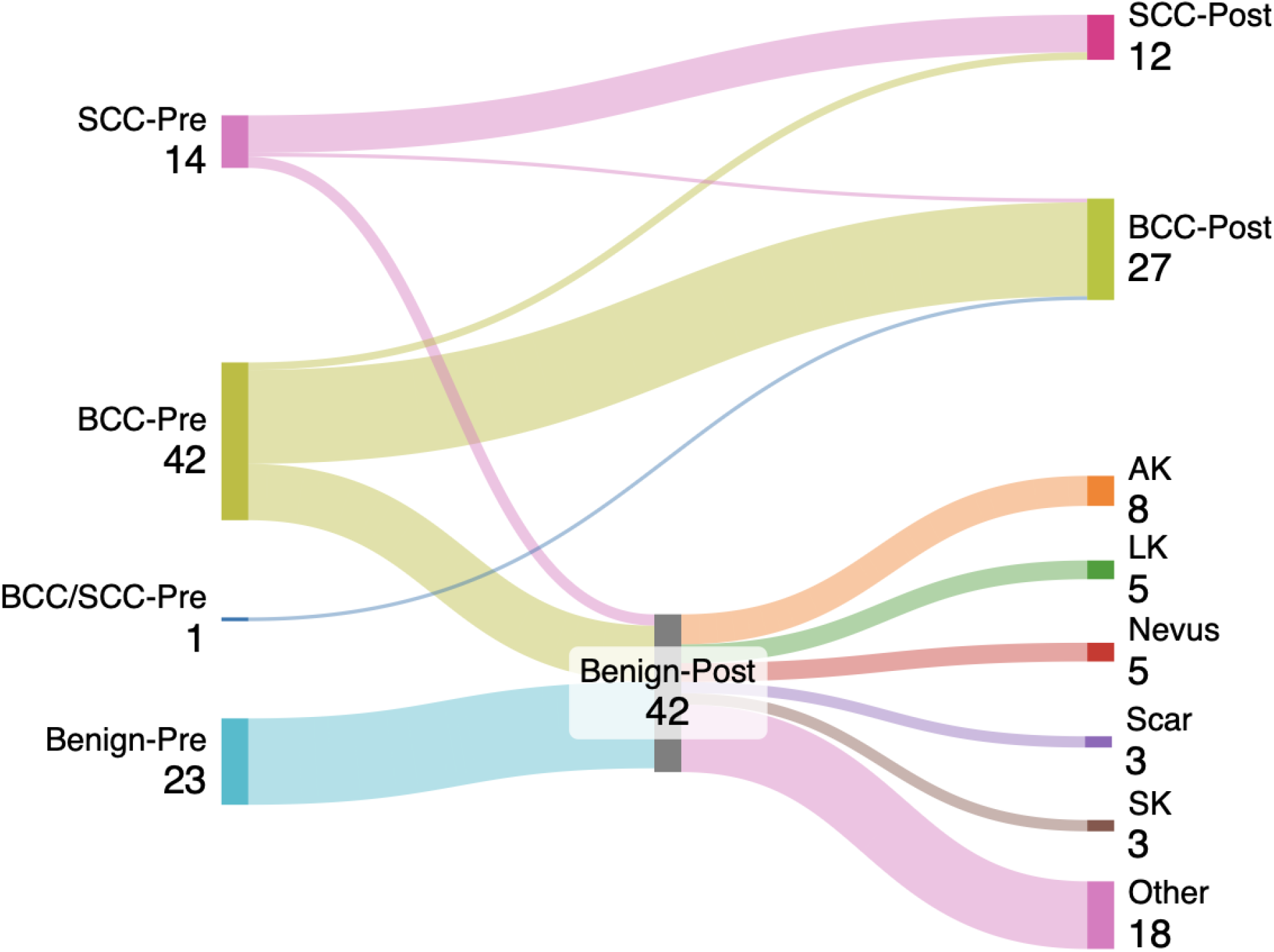
Comparison of Pre- and Post-Biopsy Diagnoses. Sankey diagram categorizing the pre-biopsy diagnoses (left) and the subsequent confirmed diagnoses (right). Out of 80 total biopsies, the resident predicted 57 to be skin cancer, while only 40 were confirmed to be skin cancer. 18 of the 57 were benign lesions. This results in a specificity of 56%.

Regarding resident exam abilities, of the 80 lesions biopsied, 40% were presented by patients as lesions of concern, while 60% were noted by a physician during physical exam. Out of the 60% that were not brought to the resident’s attention by the patient, the resident denoted that he would have biopsied 42 of the 48 lesions that the attending biopsied. 40 of those lesions were skin cancer, 36 of which the resident would have biopsied and 4 of which the resident would have missed without attending supervision. Thus, the resident correctly identified and recommended biopsy for 90% of skin cancers not brought to attention by the patient and did not identify 10%, all of which were BCCs.

## Discussion

### Overview

This observational study evaluates the sensitivity and specificity of a single second year dermatology resident in diagnosing non melanoma skin cancers. Although based on the biopsy experience of one resident, this study provides data in an area where limited information currently exists and highlighted the need for future research focused on competency-based benchmarks in dermatology residency training. In practice, studies show that dermatologists correlate clinical diagnosis and histopathological diagnosis in about 68% of cases[5]. Thus, an overall diagnostic accuracy of 64% in this analysis appears to reflect strong diagnostic abilities in early training. In addition, dermatologists were found to have the lowest NNB (2.82) when compared to dermatology advanced practice professionals (4.69), primary care physicians (4.55), and other non-dermatology clinicians (6.55)[6]. The data in this analysis yielded a NNB of 2.1, with no missed diagnoses of biopsied lesions (i.e. no lesions clinically assessed as benign that were ultimately malignant on histopathology), potentially indicating a refined biopsy use in clinic. However, there is limited published data correlating suspected clinical diagnoses with confirmed histopathologic diagnoses for residents during training, thus limiting the availability of true comparative benchmarks.

### Specificity

Specificity is important in practice due to its utility in foregoing unnecessary biopsies. In this analysis, we present a specificity of 56%. This value likely reflects a heightened level of diagnostic uncertainty early in training, resulting in biopsies of lesions that may not have otherwise required intervention. This approach is understandable during early residency training, and a goal of training would be for improved specificity over time. Notably, it is important to recognize that lower specificity early in training may be viewed as acceptable or preferable, as it may allow for fewer missed diagnoses and additionally increases trainee engagement with biopsy techniques essential for procedural education.

### Sensitivity

Sensitivity is critical in clinical decision-making, as it reflects the ability to appropriately investigate lesions concerning for true cutaneous malignancies. A goal of biopsy is to identify cutaneous malignancies before progression or spread to less accessible areas. Thus, a high sensitivity is preferred to facilitate early diagnosis. However, it is important to note that the current metastasis rate for cutaneous squamous cell carcinomas is generally estimated to be between 2-5%, with basal cell carcinomas metastasizing at an even less frequent rate[7,8].

Additionally, many of these cancers are identified on anatomically sensitive and cosmetically vulnerable areas, such as the face (30% of lesions in this analysis). With these factors in mind, patient involvement in shared decision-making prior to biopsy is imperative to balance potential risks and benefits specific to each patient.

### Limitations

A key limitation of this study is the small sample size, involving a single resident in one dermatology training program, which may limit generalizability. Additionally, details regarding clinical impressions provided to pathologists, use of diagnostic adjuncts such as dermoscopy, and resident confidence in pre-biopsy diagnosis were not recorded. These features may offer insight into factors contributing to diagnostic concordance.

### Future Directions

Future studies involving data recorded from numerous dermatology residents across multiple training programs would substantially expand this study. Nevertheless, this pilot study offers a useful framework for assessing resident diagnostic accuracy and underscores the need for further research aimed at generating evaluative standards. As dermatology residency programs across the United States shift towards competency-based curricula, this study highlights the need for developing data-driven benchmarks[9]. Greater awareness of resident’s confidence and accuracy in diagnosing keratinocyte carcinomas could inform residency guidelines, provide a baseline for comparison of practice standards across programs, and support a successful transition into independent clinical practice.

## Conclusion

Accurate and prompt diagnosis of keratinocyte carcinomas is a crucial component of dermatology residency training and frequently necessitates biopsy. In this study, one resident recorded pre- and post-biopsy diagnoses across three large academic medical centers in Boston during the second year of residency, correctly identifying the diagnosis in 64% of cases, with a specificity of 56% and NNB of 2.1. This data is a marker of diagnostic ability and is not only of value to the trainee’s longitudinal development but also for institutions seeking to evaluate and refine dermatology residency curricula. Therefore, the data presented in this study emphasizes the importance of evidence-based residency training guidelines that would enhance resident preparedness for independent practice and offer insight into variations in practice patterns.

As programs strive to update training practices with the goal of improving trainee outcomes, there is often limited information on how these updates impact clinical performance. These updates arise from evolving educational priorities within dermatology and are implemented with strong intentions. However, there is limited baseline data to assess improvement after implementation[10]. When data is collected during times of curricular change, meaningful adjustments can be incorporated, improving not only the overall education of residents but also trainee confidence as they move into more advanced levels of practice[11]. Tracking data points for residents pertaining to biopsied skin lesions with concern for malignancy would allow for evidence-based adjustments to academic programming to support the competencies already outlined by the ABD and ACGME.

Importantly, it is acknowledged that additional evaluation in graduate medical education may increase workload for both trainees and administrators. Survey fatigue can reduce trainee participation in program feedback and limit efforts for quality improvements initiatives[12,13]. Thus, more automated collection methods may minimize time burden on trainees. Additionally, trainee engagement in data collection may improve when institutions transparently share collected data and clearly communicate any data that will inform change[13].

Overall, limited data exists regarding the impact of dermatology residency curricula on pre-biopsy diagnostic accuracy. Further data collection across multiple programs and years of training would allow for improved statistical comparisons and may contribute to meaningful improvements in both residency education and patient care.

## Data Availability

All data produced in the present study are available upon reasonable request to the authors.

## Notes

**Funding sources:** JSS is supported by award K08AR084617 from the National Institute of Arthritis and Musculoskeletal and Skin Diseases.

**Conflicts of interest:** JSS is a consultant and/or investigator for Biogen and Formation Bio.

### Competing Interest Statement

JSS is a consultant and/or investigator for Biogen and Formation Bio.

### Funding Statement

JSS is supported by award K08AR084617 from the National Institute of Arthritis and Musculoskeletal and Skin Diseases.

### Summary of Updates

Manuscript revised to expand upon current analysis.

